# Invisible by Design: Three Mechanisms That Render Dementia Undetectable in Correctional Statistics Across Four High-Income Countries

**DOI:** 10.64898/2026.03.20.26348939

**Authors:** Hiroki Fukui

## Abstract

**Background:** The ageing of incarcerated populations is accelerating across high-income countries, yet dementia remains absent from routine correctional mental health statistics. We investigated whether correctional data systems in Japan, the United States, the United Kingdom, and Australia are structurally capable of detecting dementia in their prison populations.

**Methods:** We conducted a cross-national descriptive analysis of publicly available, aggregate-level correctional data. Japanese data comprised all newly admitted sentenced prisoners from 2006 to 2024 (approximately 390,000 individuals) from the Ministry of Justice Correctional Statistics Annual, including mental disorder classifications and CAPAS-derived work aptitude scores (used as a proxy for cognitive functioning; not clinical IQ measurements). US data were drawn from the Bureau of Justice Statistics Survey of Prison Inmates (2016). UK data were obtained from the Ministry of Justice Offender Management Statistics Quarterly (2015–2025). Australian data were sourced from the Australian Institute of Health and Welfare National Prisoner Health Data Collection (2022, n = 371). All analyses were descriptive; no inferential statistics were conducted.

**Findings:** Three distinct mechanisms rendered dementia statistically invisible across all four countries. First, in the United States and Australia, reliance on self-report instruments produced a paradox in which self-reported mental disorder prevalence declined with age: among US state prisoners, reported prevalence fell from 44.9% in the 35–44 age group to 31.9% among those aged 65 and older — the opposite of community epidemiological patterns. Second, in Japan — the only country with systematic cognitive assessment at prison admission — 35.0% of female theft offenders had work aptitude scores below 70, yet the classification system contains no dementia category; 43–52% of all detected mental disorders were absorbed into a residual “other” category even after a 2023 classification revision that added four new diagnostic categories but not dementia. Third, the United Kingdom lacks routine mental health prevalence data collection in prisons altogether. None of the four countries includes dementia as a standard correctional classification category.

**Interpretation:** Correctional mental health statistics across four high-income countries are structurally incapable of detecting dementia — not through clinical ignorance but by design: systems built for younger populations that have not been updated as prison demographics have changed. Japan’s ageing female theft offender profile (39.4% aged 60 or older, 35.0% with low cognitive scores) represents a potential sentinel population for undetected cognitive impairment. Targeted interventions — cognitive screening at admission in the United States and Australia, introduction of a dementia classification category in Japan, and routine mental health data publication in the United Kingdom — are feasible with existing infrastructure. As prison populations continue to age, the statistical invisibility of dementia constitutes an escalating failure of health surveillance with direct consequences for clinical care, sentencing, and human rights.

## Introduction

> *In 1793, Pinel’s act of liberation began not with a new treatment but with recognition — seeing illness where others saw only deviance. Today, correctional statistics across high-income countries lack the categories to recognise dementia in ageing prisoners at all*.

The ageing of incarcerated populations is now a defining demographic challenge for prison systems across high-income countries. In Japan, prisoners aged 50 and older constituted 41.0% of all newly admitted sentenced prisoners in 2024, making it the most aged prison population among comparable nations. The United States reported that 24.4% of state and federal prisoners were aged 50 and older in 2023, while in England and Wales the figure reached 18.2% of the total prison population in 2025. Australia, with 15.9% of its prison population aged 50 and older in 2024, shows the same upward trajectory. In all four countries, these proportions have increased steadily over the past two decades, driven by longer sentences, tougher parole policies, and demographic shifts in the general population. The epidemiological implications are substantial: community prevalence of dementia approximately doubles every five years after age 65, and accelerated biological ageing among prisoners means that age-related conditions may emerge a decade earlier than in the general population (Williams et al., 2012).

Clinical research has consistently demonstrated that dementia and cognitive impairment are present at meaningful prevalence among older prisoners. Forsyth et al. (2020, 2025) found that 7–8% of prisoners aged 50 and older in England met criteria for dementia or mild cognitive impairment in a structured clinical assessment, yet only 3% had any cognitive diagnosis recorded in their medical notes. In Germany, Verhülsdonk et al. (2020) reported cognitive dysfunction in 41.4% of older prisoners using standardised neuropsychological testing. Combalbert et al. (2018) found moderate-to-severe cognitive impairment in approximately 20% of older prisoners in France. These studies leave little doubt that dementia exists within prison walls at rates that demand systematic attention.

Yet this clinical evidence has not penetrated the statistical infrastructure through which correctional systems understand their populations. The largest systematic reviews of prisoner mental health — Fazel and Baillargeon (2011) in the Lancet and, more recently, Emilian and Fazel (2025) in a 43-country meta-analysis — did not include dementia among the conditions examined. This omission is not incidental; it reflects the structure of the data available. If national correctional statistics do not classify dementia, meta-analyses drawing on those statistics cannot find it. The question, then, is not simply whether prisoners have dementia — clinical research has answered that — but whether the statistical systems designed to monitor prisoner health have the capacity to detect it at all.

This paper asks whether correctional mental health statistics in four high-income countries — Japan, the United States, the United Kingdom, and Australia — are structurally capable of rendering dementia visible. Through a cross-national analysis of routine correctional data, we identify three distinct mechanisms by which dementia is excluded from statistical view: the reliance on self-report instruments that cognitively impaired individuals cannot reliably complete (United States, Australia), the absence of a dementia category in classification systems that otherwise measure cognitive functioning (Japan), and the failure to collect routine mental health prevalence data altogether (United Kingdom). These three mechanisms are not separate problems in separate countries; they are three manifestations of a single structural condition — correctional statistics, as currently designed, cannot see dementia. We use the term ‘by design’ not to imply deliberate intent to exclude dementia, but to describe a structural condition: systems built for younger populations, using instruments chosen before prison populations began to age, that have not been updated as the epidemiological reality has changed.

## Methods

### Data Sources and Study Design

This study employed a cross-national descriptive analysis of routine correctional mental health data from four high-income countries: Japan, the United States, the United Kingdom, and Australia. No individual-level linked datasets were available; all analyses were conducted on aggregate, population-level statistics published by government agencies. We examined the structure of mental health classification systems, the categories available for recording cognitive and psychiatric conditions, and the patterns of detection across age groups and offence types. The study did not involve primary data collection or direct contact with incarcerated individuals.

### Japanese Correctional Data

Japanese data were drawn from the Correctional Statistics Annual (*Ky*◼*sei T*◼*kei Nenp*◼), published by the Ministry of Justice and accessed via the e-Stat portal (https://www.e-stat.go.jp/). The dataset covers all newly admitted sentenced prisoners (*shin-ju-keisha*) from 2006 to 2024, comprising approximately 390,000 individuals over the study period (female: n = 35,082; male: n = 355,353). These are census data, not samples: every newly admitted prisoner is included.

At reception, each prisoner undergoes a screening assessment conducted by correctional facility staff (non-specialist), with referral to physicians when clinically indicated. This process yields two types of data relevant to the present analysis:

1. **Mental disorder classification**. Prior to 2023, prisoners were classified into five categories: no mental disorder, intellectual disability, personality disorder, neurotic disorder, and other mental disorder. In 2023, the classification was expanded to nine categories, adding developmental disorder, substance use disorder, schizophrenia, and mood disorder. Critically, no version of the classification has included a category for dementia or cognitive impairment (i.e., no equivalent to ICD-10 codes F00–F03 or G30).
2. **Work aptitude assessment (CAPAS)**. Japan is unique among the four countries in administering a standardised work aptitude test to all prisoners at admission. The CAPAS (*Correctional Association Psychological Assessment Series*) is designed to assess work aptitude for prison labour allocation. The CAPAS yields IQ-equivalent values as a by-product of its work aptitude assessment; we refer to these hereafter as work aptitude scores (WAS), acknowledging that they are not validated intelligence measurements. We report WAS scores as a descriptive indicator of cognitive functioning at the population level.

### International Comparative Data

#### United States

United States data were obtained from the Bureau of Justice Statistics (BJS) Survey of Prison Inmates (SPI), 2016. The SPI is a nationally representative cross-sectional survey of state and federal prisoners. Mental health status was assessed exclusively through self-report: prisoners reported whether they had ever been told by a mental health professional that they had any of six disorder categories (major depressive disorder, bipolar disorder, anxiety disorder, PTSD, personality disorder, or schizophrenia/psychotic disorder). The Kessler-6 (K6) screening scale was also administered to measure serious psychological distress (SPD). No cognitive testing, WAS assessment, or dementia screening was conducted.

#### United Kingdom

United Kingdom data on prison demographics were obtained from the Ministry of Justice (MOJ) Offender Management Statistics Quarterly (OMSQ), covering the total prison population from 2015 to 2025. Mental health clinical data in England and Wales are held by the National Health Service (NHS) in electronic health records (SystmOne), using ICD-10/ICD-11 coding. However, these clinical data are not routinely published as correctional statistics, creating a structural separation between demographic and health information. No routine cognitive screening or dementia assessment is conducted at prison reception. Published research on prison dementia prevalence exists (notably Forsyth et al., 2020, reporting 8% dementia/MCI prevalence among prisoners aged 50 and older, n = 869), but these findings are not integrated into routine surveillance.

#### Australia

Australian data were drawn from two sources: the Australian Institute of Health and Welfare (AIHW) National Prisoner Health Data Collection (NPHDC), 2022, and the Australian Bureau of Statistics (ABS) Prisoners in Australia series. The NPHDC is a biennial cross-sectional survey conducted at prison reception. The 2022 cycle included a voluntary sample of 371 prison entrants and 431 dischargees. Mental health status was assessed through a single self-report item (“Have you ever been told by a doctor, psychiatrist, psychologist or nurse that you have a mental health condition?”) supplemented by the Kessler-10 (K10) psychological distress scale and data on psychotropic medication use. No cognitive screening, WAS assessment, or dementia assessment was performed. The small sample size (n = 371 for entrants) limits generalisability.

### Measures

The primary outcomes examined were:

- **Mental disorder detection rate**: the proportion of newly admitted prisoners identified as having any mental disorder (Japan).
- **Mental health prevalence indicators**: self-reported history of mental health conditions and K6/K10 distress scores (US, Australia).
- **Classification structure**: the presence or absence of specific diagnostic categories, particularly dementia and cognitive impairment, in each country’s data system.
- **Age distribution**: the proportion of prisoners aged 50 and older (or 55+/60+, depending on data availability) as an indicator of population ageing.
- **Work aptitude scores (WAS)**: CAPAS-derived scores categorised as <50, 50–69, and ^3^70, reported as a proxy for cognitive functioning (Japan only).
- **Offence type**: principal offence at admission, with particular attention to theft offences among older prisoners.

### Analytical Approach

All analyses were descriptive. We calculated proportions, trends over time, and cross-tabulations of mental health indicators by age, sex, and offence type. No inferential statistics or hypothesis tests were conducted. For Japanese data, we computed annual detection rates and category-specific proportions across the 2006–2024 period. For the US self-report paradox analysis, we compared mental health indicator prevalence across age groups within the 2016 SPI cross-section. International comparisons were necessarily qualitative given differences in assessment methods, target populations, time periods, and sample frames across the four countries (Table 3).

### Ethical Considerations

This study used only publicly available, aggregate-level administrative data. No individual-level data were accessed, and no data linkage was performed. The Japanese correctional data are published as official statistics through e-Stat with no access restrictions. The US BJS data are publicly available summary tables. UK MOJ data are published quarterly. Australian AIHW data are publicly available reports. As the study involved secondary analysis of de-identified, aggregate data, institutional ethics review was not required.

## Results

### Section A: Japanese Correctional Data

#### Ageing of the prison population

The proportion of older prisoners among newly admitted individuals in Japan increased substantially over the study period (Fig 1). Prisoners aged 50 and older rose from 29.4% in 2006 to 41.0% in 2024, while those aged 60 and older increased from 11.3% to 20.4% over the same period. Japan had by far the highest proportion of older prisoners among the four countries examined: by comparison, the UK total prison population was 18.2% aged 50 and older in 2025, the US state and federal prison population was 24.4% aged 50 and older in 2023, and Australia reported 15.9% aged 50 and older in 2024 (Fig 1).

**Fig 1.**
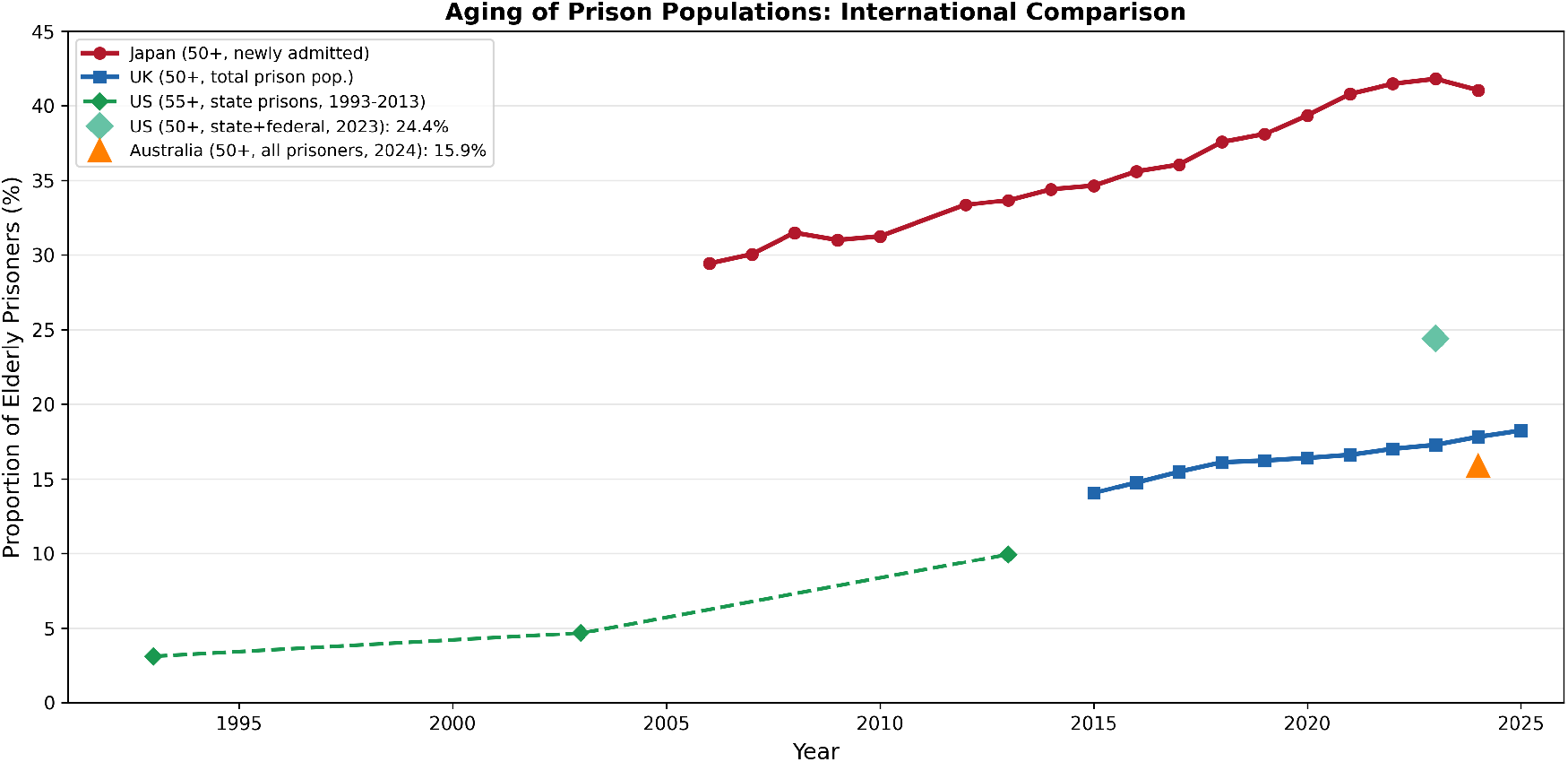

#### Rising mental disorder detection rates

The overall detection rate for mental disorders among newly admitted prisoners rose markedly across the study period (Fig 4). Among females, the detection rate increased from 14.3% in 2006 to 35.3% in 2024—a 2.5-fold increase. Among males, the rate rose from 4.8% in 2006 to 20.6% in 2024—a 4.3-fold increase. Both trends showed near-monotonic increases, with the steepest acceleration occurring after 2022.

#### Diagnostic category distribution and the dominance of “Other”

Prior to the 2023 classification revision, the “Other mental disorder” category absorbed the majority of all detected cases across both sexes (Fig 5A, Fig 5B). Among females, “Other” accounted for 64.3%–81.9% of all detected mental disorders between 2006 and 2022. Among males, “Other” accounted for 55.2%–73.1% of detected cases over the same period.

The 2023 classification revision, which added four new diagnostic categories (developmental disorder, substance use disorder, schizophrenia, and mood disorder), partially reduced the dominance of “Other.” Among females, “Other” dropped from 78.3% in 2022 to 47.1% in 2023 and 43.3% in 2024. Among males, “Other” dropped from 67.9% in 2022 to 49.1% in 2023, before rising slightly to 52.5% in 2024.

Notably, even after the 2023 revision, the “Other” category continued to absorb approximately half of all detected cases. Furthermore, the 2023 revision added categories for schizophrenia, mood disorder, developmental disorder, and substance use disorder, but did not add a category for dementia or cognitive impairment—despite the concurrent ageing of the prison population (Fig 5A, Fig 5B). (Note: The 2023 classification revision introduced four new categories — developmental disorder, substance use disorder, schizophrenia, and mood disorder. These are not separately represented in the publicly available aggregate data used for these figures; their introduction accounts for the observed decline in the ‘Other Mental Disorder’ proportion from 2023 onwards, as cases previously absorbed into ‘Other’ were redistributed to the new specific categories.)

#### Female theft offender profile

Female theft offenders presented a distinctive demographic and cognitive profile (Table 1, Fig 7). Among female theft offenders (n = 14,867), 39.4% were aged 60 or older—more than five times the rate among female non-theft offenders (7.6%). The rate of WAS scores below 70 was 35.0% among female theft offenders, compared with 14.8% among female non-theft offenders. The mental disorder detection rate was 23.2% for female theft offenders versus 22.2% for female non-theft offenders.

Among male prisoners, a parallel but less extreme pattern was observed: 23.2% of male theft offenders were aged 60 or older compared with 12.3% of male non-theft offenders. WAS scores below 70 were found in 28.7% of male theft offenders versus 17.0% of male non-theft offenders (Table 1).

#### The work aptitude score–diagnosis gap

A striking discrepancy emerged between cognitive test performance and diagnostic classification (Table 2). Among female theft offenders, 35.0% had WAS scores below 70 (8.4% below 50, 26.7% in the 50–69 range), yet only 1.1% received a diagnosis of intellectual disability. Among female prisoners overall, 23.4% scored below 70, while 0.8% were diagnosed with intellectual disability. The “Other mental disorder” category accounted for 14.9% of female theft offenders, suggesting that cognitively impaired individuals were either classified under this residual category or not captured by any diagnostic label.

Among males, the gap was similarly pronounced: 28.7% of male theft offenders had WAS scores below 70, compared with a 2.1% intellectual disability diagnosis rate. For males across all crime types, 20.9% scored below 70 versus 1.3% diagnosed with intellectual disability (Table 2).

### Section B: International Comparison

#### Data structure comparison across four countries

The four countries differed fundamentally in how they collected and classified prison mental health data (Table 3). Japan was the only country employing clinician-based assessment at reception and the only country routinely measuring cognitive ability. The United States and Australia relied entirely on self-report. The United Kingdom collected clinical data through the NHS but did not routinely publish it as correctional statistics. None of the four countries included dementia or cognitive impairment as a category in their routine prison mental health classification systems. The US and Australian systems lacked any cognitive or intellectual disability assessment. Japan’s system lacked a dementia category despite being the only country with systematic cognitive measurement. The UK had research-based prevalence estimates (Forsyth et al., 2020) but no routine screening programme (Table 3).

#### The self-report paradox in US prison data

Analysis of the 2016 BJS Survey of Prison Inmates revealed a counterintuitive age-related pattern in mental health reporting among state prisoners (Fig 2). Self-reported history of any mental health disorder peaked at 44.9% among prisoners aged 35–44, then declined progressively to 42.8% (aged 45–54), 38.4% (aged 55–64), and 31.9% (aged 65 and older)—a 13.0 percentage point decline from peak to the oldest age group. The K6 serious psychological distress measure showed a parallel pattern: SPD prevalence was 15.0% among those aged 35–44 but fell to 9.9% among those aged 65 and older.

**Fig 2.**
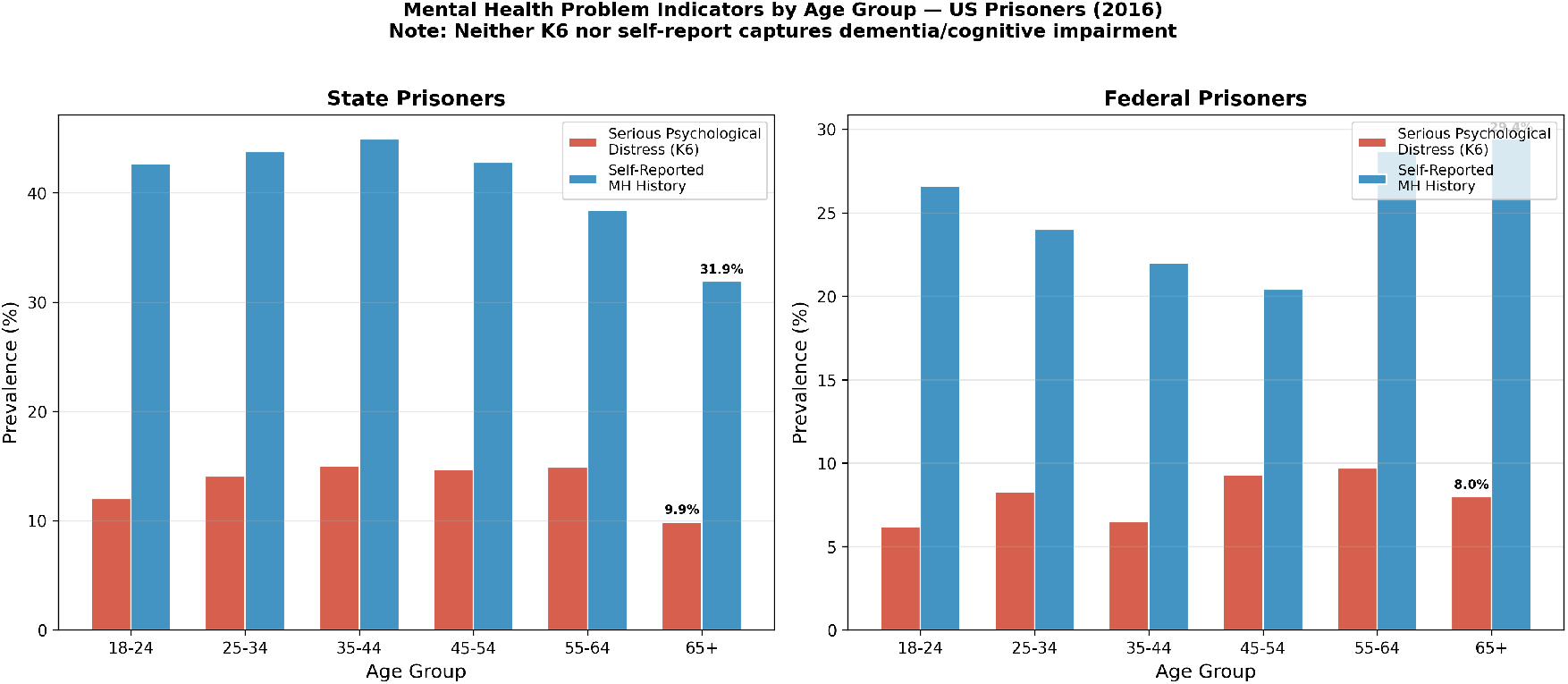

The proportion of prisoners with “no indication” of mental health problems (neither self-reported history nor elevated K6 scores) rose from 42.1% in the 35–44 age group to 56.4% in the 65 and older group—a 14.3 percentage point increase (Fig 2). This pattern—in which the oldest prisoners appear to be the healthiest on mental health measures—runs counter to community epidemiological evidence showing increasing prevalence of mental health conditions and cognitive impairment with age. (Note: Panel A of Fig 3 presents the US state prisoner data from this figure with annotated age zones for comparison with international data.)

**Fig 3.**
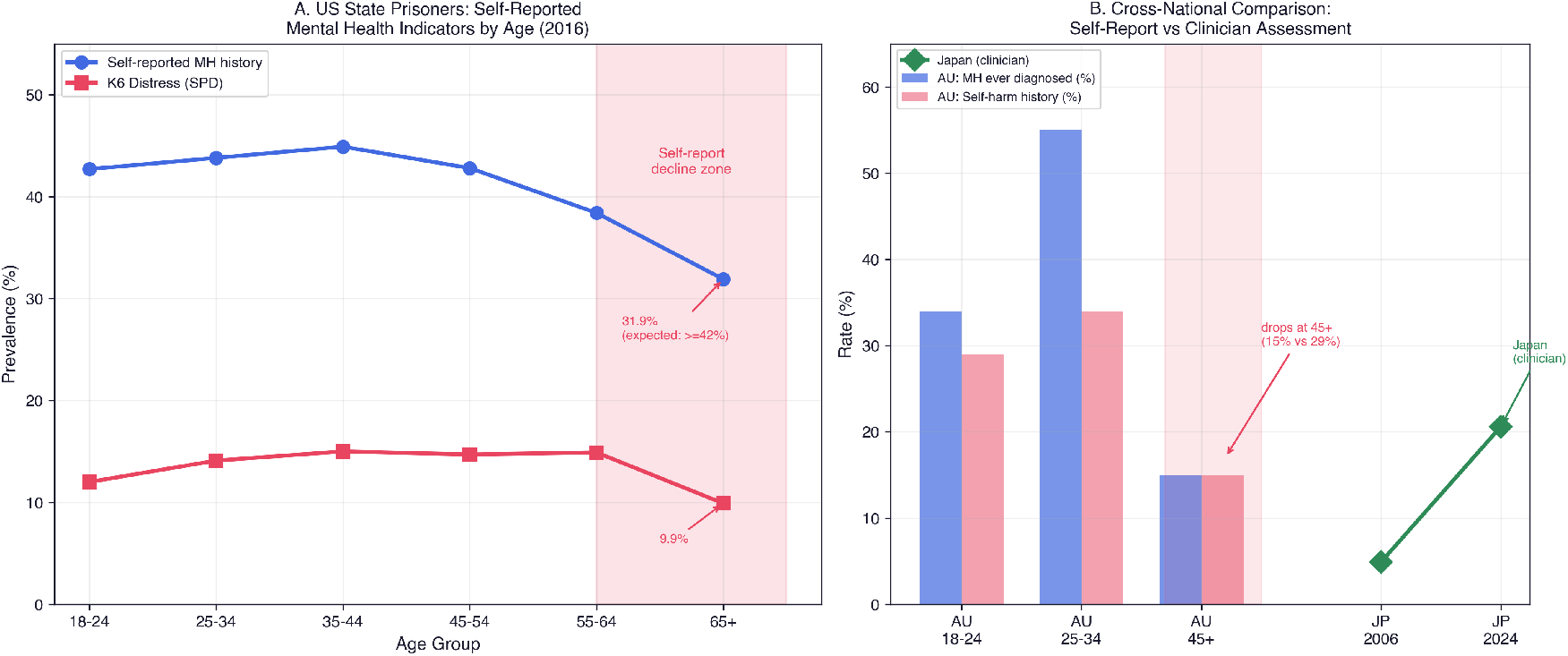

**Fig 4.**
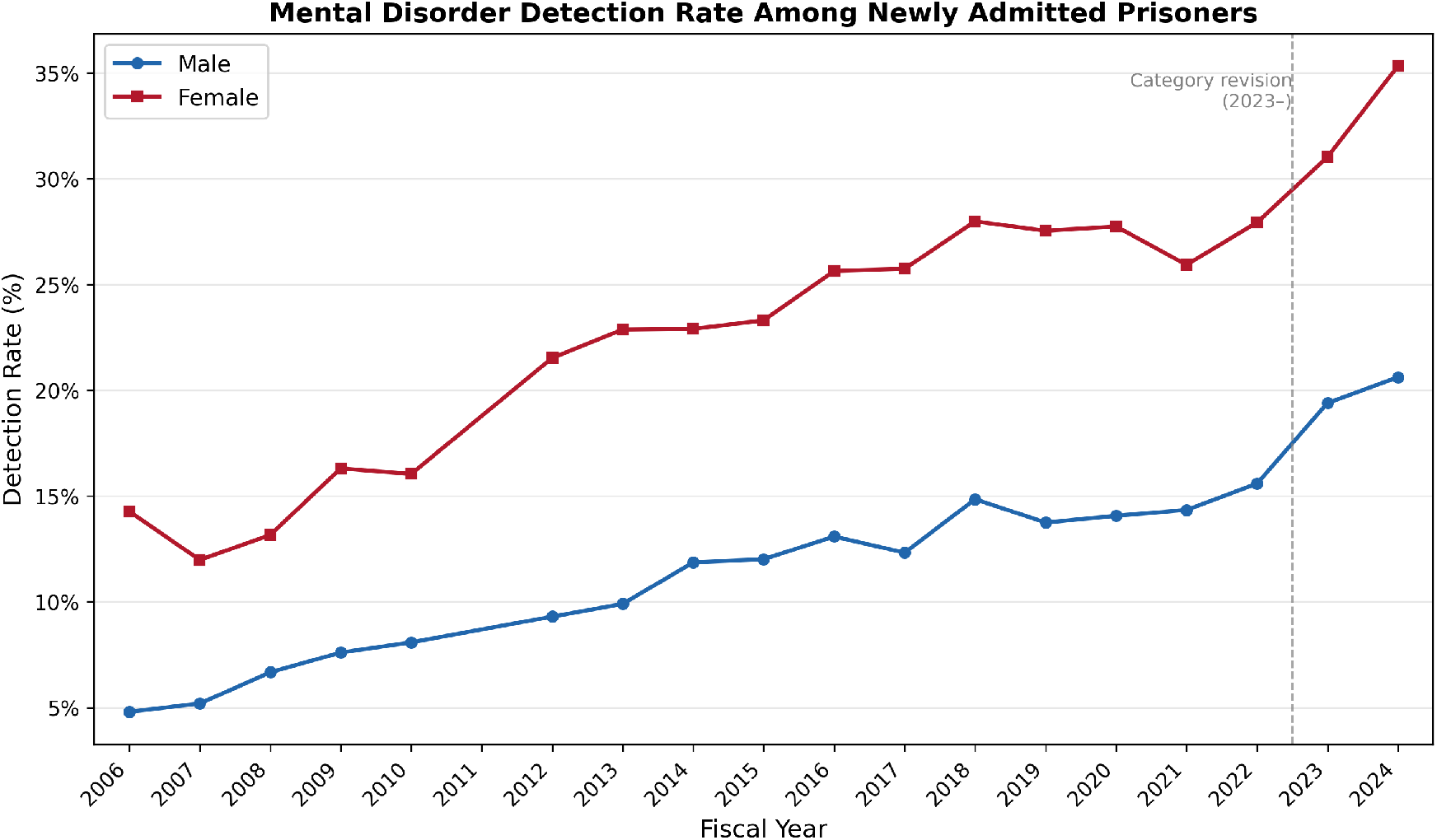

**Fig 5A.**
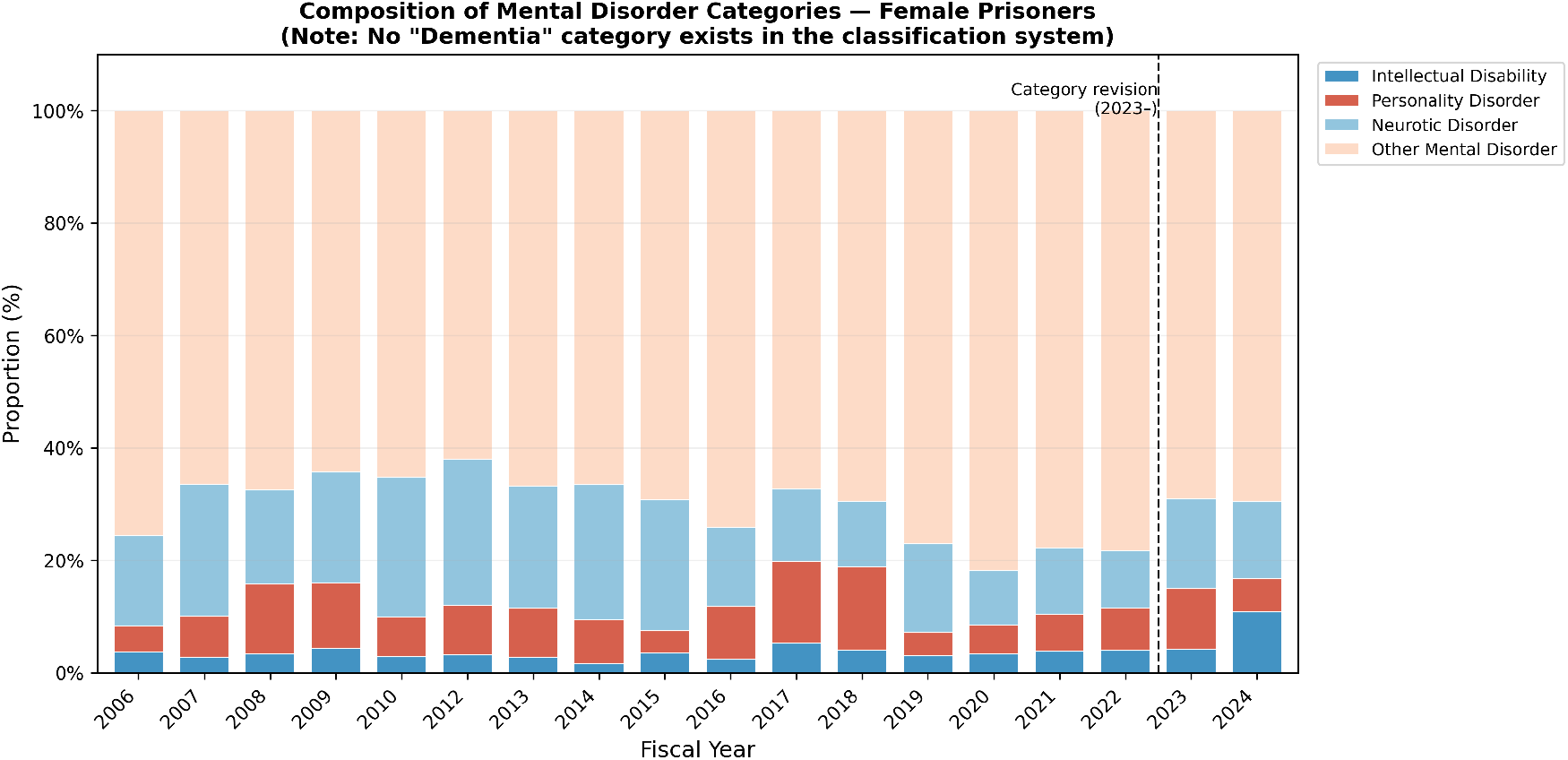

**Fig 5B.**
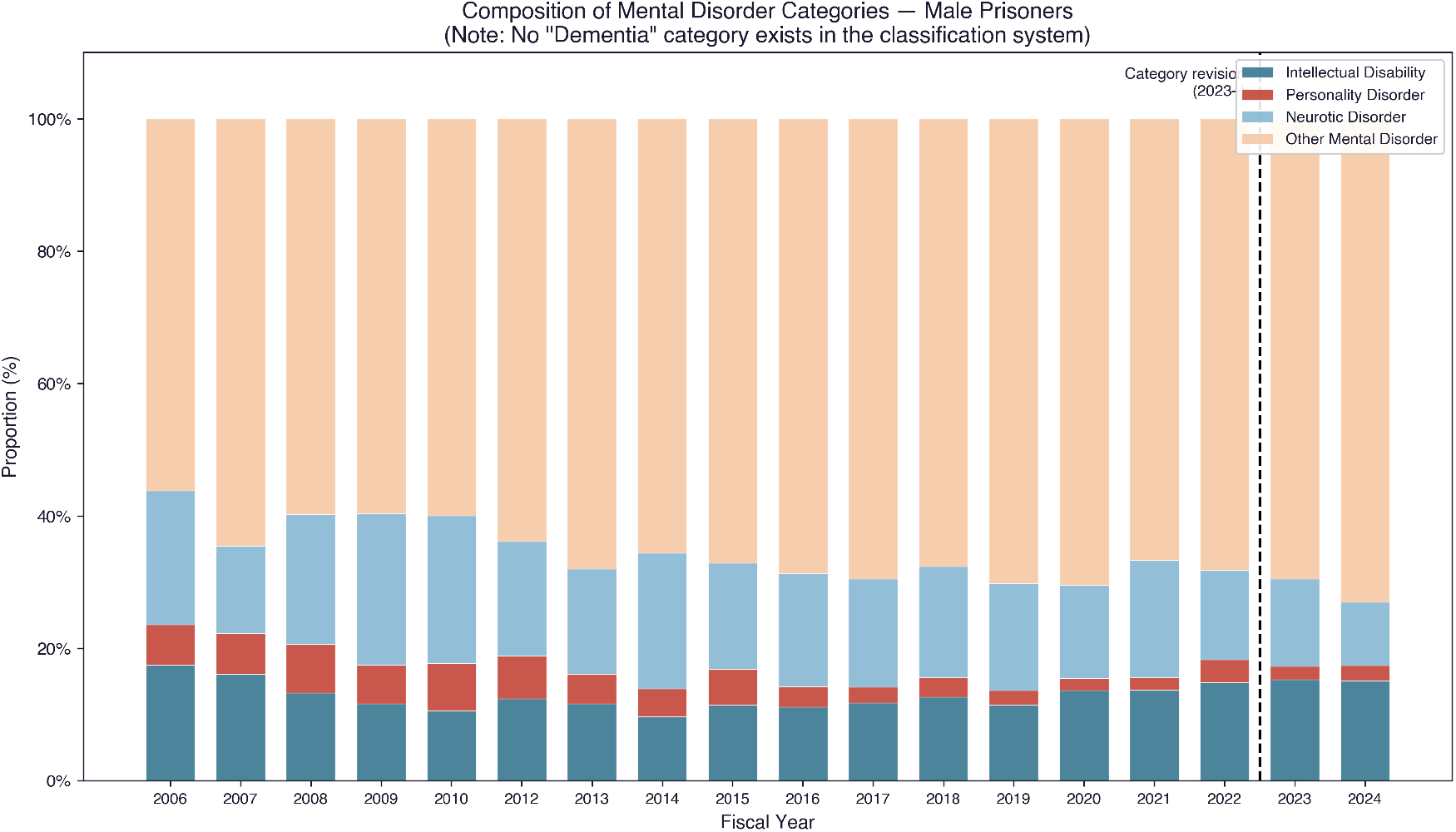

#### International confirmation of the paradox

The self-report paradox in context — annotated US data and international confirmation — is presented in Fig 3. The self-report paradox observed in US data was partially confirmed in Australian data (Fig 3). In the 2022 AIHW NPHDC, self-harm history declined from 29% among entrants aged 18–24 to 15% among those aged 45 and older. Self-harm ideation in the prior 12 months decreased from 20% (aged 25–34) to 9.8% (aged 45 and older). These self-report-based indicators showed the same directional pattern as US data: older prisoners reported fewer mental health difficulties despite being in a demographic group with higher expected prevalence of cognitive conditions (Fig 3).

Japan’s clinician-assessed system showed the opposite trajectory: mental disorder detection rates increased alongside population ageing, with detection rates rising monotonically alongside population ageing across the entire study period (Fig 3). This divergence between self-report systems (declining indicators in older groups) and clinician-assessed systems (rising detection in ageing populations) highlights the methodological sensitivity of mental health measurement in correctional settings.

#### Offence profiles by age: Japan–Australia contrast

The relationship between age and offence type differed markedly between Japan and Australia (Fig 6). In Japan, ageing was strongly associated with theft offences: 39.4% of female theft offenders and 23.2% of male theft offenders were aged 60 or older, compared with 7.6% and 12.3% of non-theft offenders, respectively (Table 1). In Australia, by contrast, the overrepresentation of sexual offences among older prisoners has been documented in the literature (Crookes et al., 2022). This divergence in offence profiles has implications for the types of cognitive impairment likely to be present—and missed—in each country’s ageing prison population (Fig 6).

**Fig 6.**
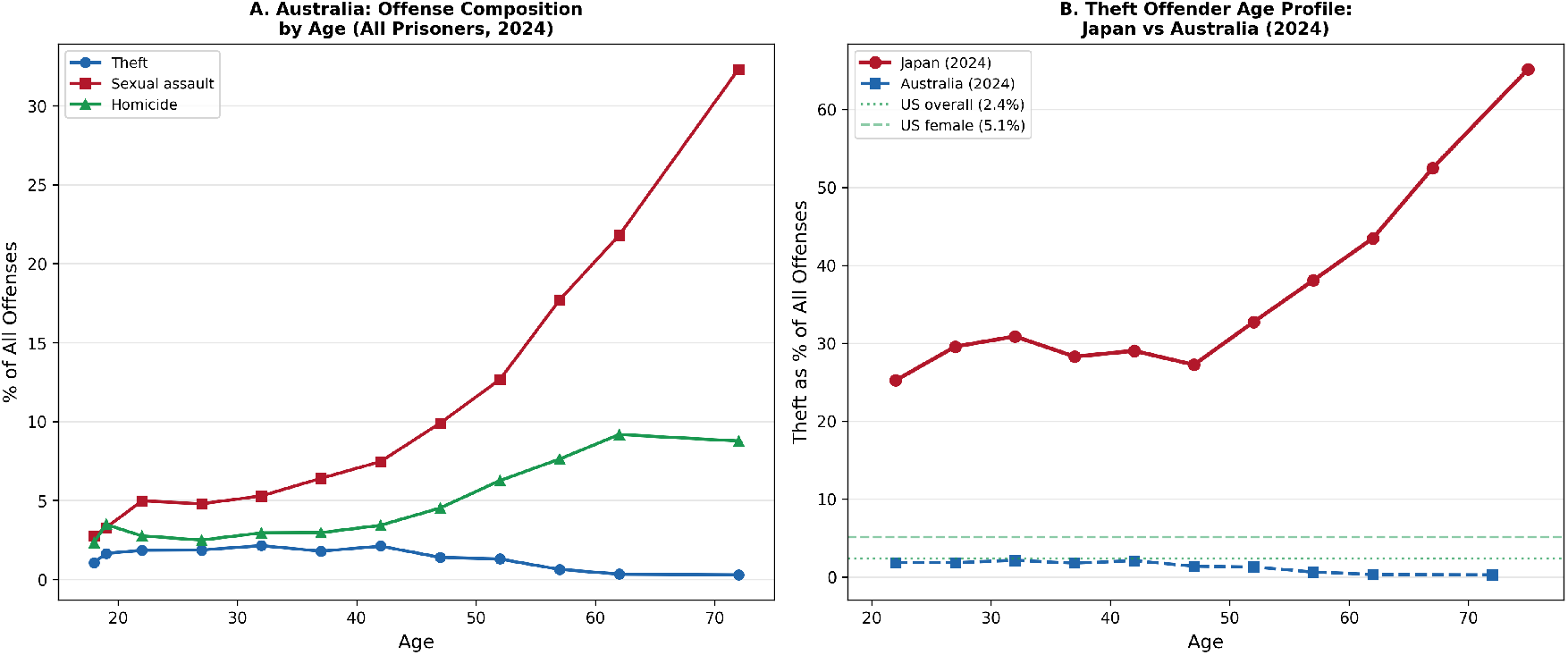

**Fig 7.**
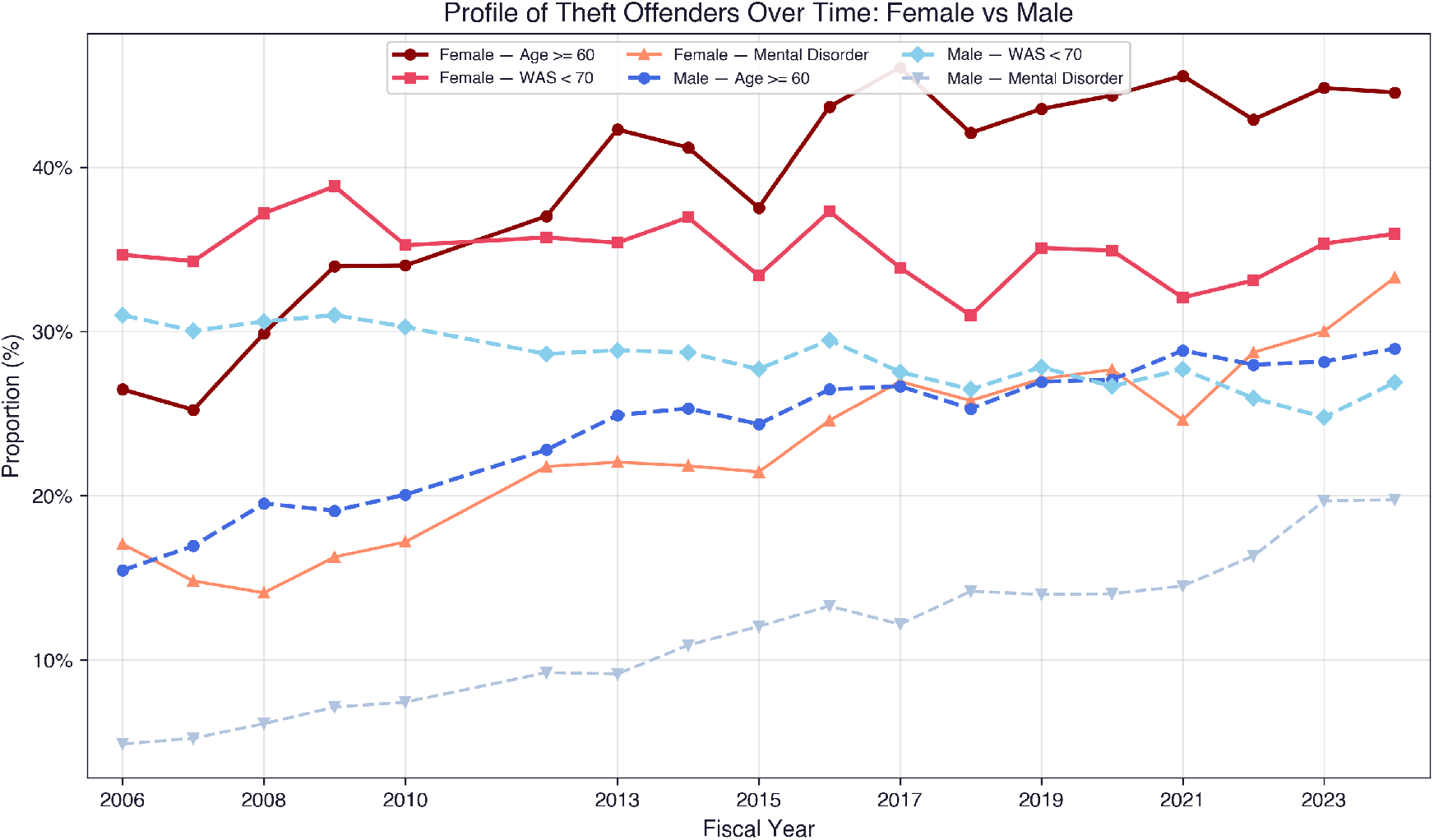

*Note: All analyses are descriptive. No inferential statistical tests were conducted. Direct comparison of prevalence rates across countries is not possible due to differences in assessment methods (clinician-based vs self-report), target populations (newly admitted vs total population vs sample), time periods, and sample sizes. The CAPAS work aptitude scores are reported as descriptive indicators of cognitive functioning and should not be interpreted as clinical IQ measurements*.

## Discussion

### Principal findings

This cross-national analysis of correctional mental health data from Japan, the United States, the United Kingdom, and Australia reveals that the absence of dementia from prison statistics is not an oversight in any single country but a structural condition embedded in how correctional systems collect, classify, and report health information. We identified three distinct mechanisms through which dementia is rendered statistically invisible, each representing a different manifestation of the same underlying problem: correctional data systems, as currently designed, lack the capacity to see dementia. In the United States and Australia, the reliance on self-report instruments means that the very population most likely to have cognitive impairment is the least able to report it. In Japan, cognitive functioning is measured at scale, yet the classification system contains no category into which dementia can be placed, causing detected impairment to be absorbed into residual categories. In the United Kingdom, the infrastructure for routine mental health prevalence monitoring in prisons does not exist, rendering dementia — along with much else — invisible by default.

### The self-report paradox

The age-related decline in self-reported mental health problems observed in US prison data is, on its face, implausible. Among state prisoners, the proportion reporting any mental health history fell from 44.9% in the 35–44 age group to 31.9% among those aged 65 and older — a 13 percentage point decline. The K6 serious psychological distress measure showed a parallel drop, from 15.0% to 9.9%. Community epidemiological data consistently show that the prevalence of cognitive impairment and dementia increases sharply with age; the notion that the oldest prisoners are mentally the healthiest is epidemiologically untenable.

Australian data, though derived from a smaller and methodologically different survey, confirm the same directional pattern. Self-harm history declined from 29% in the youngest entrants to 15% among those aged 45 and older, and self-harm ideation fell from 20% to under 10%. The consistency of this pattern across two countries with independent data collection systems, different instruments, and different prison populations strengthens the inference that the decline reflects a systematic measurement artefact rather than a true epidemiological trend.

Three alternative explanations for this age-related decline warrant consideration. First, healthy survivor effects: if prisoners with severe mental illness die or are released disproportionately before reaching older age groups, the surviving older population would appear healthier through selection alone. Second, cohort effects: older prisoners may belong to generations with stronger stigma around mental health disclosure, producing lower self-report rates independent of true prevalence. Third, differential incarceration patterns: offence profiles and sentencing practices differ by age cohort in ways that may confound cross-sectional mental health comparisons.

We cannot exclude these explanations with aggregate-level data alone, and we do not claim to do so. What we can observe is a convergence of three independent patterns that, taken together, are more consistent with a structural measurement artefact than with any single alternative explanation. First, the age-related decline in self-report rates occurs specifically in both the United States and Australia — two countries with independent data systems, different instruments, and different prison populations — suggesting it is not a country-specific confound. Second, the sharpest decline occurs at 65 and older, precisely the age at which dementia prevalence rises steeply in community populations; a pattern driven primarily by healthy survivor effects or cohort stigma would not be expected to cluster at this specific threshold. The more gradual decline observed between ages 35–44 and 55–64 is consistent with the alternative explanations we have noted — cohort effects and differential incarceration patterns — and we do not attribute this portion of the trend to dementia. Our claim is more specific: the acceleration of the decline at 65 and older, at precisely the threshold where community dementia prevalence rises steeply, is the pattern that most strongly implicates a measurement artefact related to cognitive impairment. Third, Japan’s clinician-assessed system — which measures the same ageing phenomenon — shows the opposite directional pattern, with detection rates rising monotonically over the same period. No single alternative hypothesis accounts for all three observations simultaneously.

The most parsimonious explanation is that self-report instruments are structurally unable to capture conditions that compromise the capacity for self-report. Dementia, by its nature, impairs the very cognitive functions — insight, memory, and executive functioning — that self-report requires. Kingston et al. (2011) have shown that cognitive impairment among older prisoners is poorly detected by self-report methods, and the K6 and K10 screening instruments were designed to measure psychological distress, not cognitive decline. This creates a paradox in which the measurement system produces the appearance of declining mental health burden precisely as the true burden is rising. The contrast with Japan’s clinician-assessed system, where mental disorder detection rates rose monotonically over the same period, underscores that the paradox is a property of the method, not of the population.

### The classification gap

Japan presents a paradox of a different kind. It is the only country among the four that routinely administers a standardised cognitive assessment to all prisoners at admission, and its data therefore offer an unusually detailed picture of cognitive functioning at the population level. The CAPAS assessment reveals that 35.0% of female theft offenders and 28.7% of male theft offenders have WAS scores below 70 — figures that, in community settings, would trigger immediate diagnostic evaluation. Yet the diagnostic classification system into which these individuals are sorted contains no category for dementia or cognitive impairment. The result is that intellectual disability — the only category that could absorb low cognitive scores — captures just 1.1% of female theft offenders, while the residual “Other mental disorder” category swells to account for 43–52% of all detected cases even after the 2023 classification revision.

The 2023 revision of Japan’s classification system is instructive. The addition of four new categories — developmental disorder, substance use disorder, schizophrenia, and mood disorder — reduced the proportion of cases classified as “Other,” but the revision did not include dementia. This was a deliberate design choice made in the context of a prison population in which over 40% of new admissions were aged 50 or older. The classification gap is therefore not a legacy problem inherited from an earlier era; it is a contemporary decision that shapes what can and cannot be counted.

The consequences are visible in the data. Japan collects more cognitive data than any other country in this analysis, yet it cannot translate that data into diagnostic categories that would make dementia visible. Detection without classification produces information that cannot be acted upon: a low CAPAS score triggers a work assignment decision, but not a clinical referral, because the system contains no pathway from cognitive test performance to dementia evaluation. The Japanese case thus illustrates that data collection alone is insufficient; what matters equally is whether the categories exist to give the data clinical meaning.

### The female theft offender as sentinel population

The profile of female theft offenders in Japan — 39.4% aged 60 or older, 35.0% with WAS scores below 70 — suggests a convergence of demographic and cognitive indicators consistent with elevated rates of cognitive impairment. The neurological literature on frontotemporal dementia (FTD) provides a plausible mechanism. Liljegren et al. (2015) reported criminal behaviour in 37.4% of patients with the behavioural variant of FTD, with theft being among the most common offence types. Mendez (2010) documented similar patterns of socially inappropriate behaviour and legal violations in FTD patients who had no prior criminal history. In Japan specifically, Shinagawa et al. (2017) found that legal violations, including shoplifting, were a recognised presentation of FTD in a multicenter study. These studies were conducted in clinical, not correctional, populations; direct evidence of FTD or dementia as a driver of offending among Japanese prisoners is absent from the literature. The inference from population-level patterns to individual-level mechanisms requires caution.

The Japan–Australia contrast in offence profiles among older prisoners sharpens this interpretation. In Australia, the overrepresentation of sexual offences among older prisoners has been documented in the literature (Crookes et al., 2022) — offences that are typically planned over extended periods and that do not, by their nature, suggest acute cognitive impairment at the time of the offence. In Japan, older female prisoners are disproportionately convicted of theft — an impulsive offence type consistent with the disinhibition and poor judgement characteristic of FTD and other dementias. This divergence suggests that Japan’s ageing female theft offender population may function as a sentinel group for undetected dementia — a population in which cognitive impairment is not merely a comorbidity but a potential driver of the offending behaviour itself. An alternative explanation — that economic hardship among older women in Japan drives shoplifting behaviour independently of cognitive impairment — cannot be excluded and likely contributes to the observed pattern. Japan’s relative poverty rate among adults aged 65 and older exceeds 20%, among the highest in the OECD, and is substantially higher for women living alone (OECD, 2023). Economic marginalisation and cognitive impairment are not mutually exclusive explanations; both may concentrate in this demographic group simultaneously, and disentangling their relative contributions requires individual-level clinical investigation that the present aggregate data cannot provide.

### Data absence

The United Kingdom illustrates a third mechanism of invisibility: the absence of routine data collection on mental health prevalence in prisons. The research of Forsyth et al. (2020, 2025) demonstrates that structured clinical assessment can identify dementia and mild cognitive impairment in 7–8% of prisoners aged 50 and older — yet only 3% of those individuals had any cognitive diagnosis recorded in their medical notes. This research exists as an isolated study, not as part of routine surveillance. The National Audit Office (2017) stated plainly that the government does not know how many people in prison have a mental illness, a conclusion that remains essentially unchanged.

Following the transfer of prison healthcare commissioning to NHS England in 2013, clinical data collected by prison healthcare teams are recorded in the SystmOne electronic health record system and governed by NHS Information Governance frameworks. These frameworks were not designed with population-level correctional surveillance in mind, and no mechanism exists for the routine aggregation and publication of prison mental health prevalence data as correctional statistics. The structural separation between NHS clinical data and Ministry of Justice correctional statistics means that even where clinical data are collected, they are not routinely available for population-level monitoring. The problem in the UK is therefore prior to the question of self-report validity or classification adequacy — it is the absence of the data infrastructure itself. Without routine collection and publication of prison mental health data, the prevalence of dementia cannot be tracked, trends cannot be identified, and the scale of the problem remains unknown.

### Dialogue with prior literature

The findings of this study must be understood in relation to the existing evidence base on prisoner mental health. Emilian and Fazel (2025) conducted the largest meta-analysis to date, synthesising data from 43 countries, yet did not include dementia among the conditions examined. This exclusion is not a limitation of the review itself but rather a reflection of the data landscape: if national correctional statistics do not classify dementia, systematic reviews cannot aggregate what does not exist. Our analysis complements this work by shifting the focus from clinical prevalence — which clinical studies have established — to the structural capacity of statistical systems to detect and record that prevalence. Brooke et al. (2020), in their review of dementia in prison settings, documented the challenges of identification and care but did not examine the data infrastructure through which detection might be systematically enabled or prevented.

This study also addresses a gap in the English-language literature on Japanese corrections. Suzuki and Otani (2023) provided the only prior English-language overview of ageing in Japanese prisons but offered primarily qualitative description without quantitative analysis of correctional data. The present study introduces Japanese correctional statistics — including the CAPAS cognitive assessment data and diagnostic classification system — to the international literature for the first time, enabling direct structural comparison with US, UK, and Australian systems. The Japanese case is particularly valuable because it demonstrates that cognitive measurement alone does not guarantee cognitive visibility; the classification framework determines what becomes actionable knowledge.

### Policy implications

The three distinct mechanisms identified in this study suggest differentiated policy responses. For countries relying on self-report instruments — currently the United States and Australia — the most immediate intervention is the introduction of brief cognitive screening for prisoners above a threshold age. Instruments such as the Montreal Cognitive Assessment (MoCA) require minimal training and can be administered in 10–15 minutes. A policy requiring cognitive screening for all prisoners aged 50 and older at admission would address the self-report paradox by supplementing subjective measures with an objective assessment of cognitive function. This is not a call to replace existing mental health screening but to add a cognitive dimension that current instruments do not capture.

For Japan, where cognitive data already exist, the priority is to introduce a dementia category into the diagnostic classification system and to establish a clinical referral pathway for prisoners with low CAPAS scores. A two-stage screening protocol — in which CAPAS scores below a threshold value trigger a structured clinical assessment for dementia — would make use of data that are already being collected. For the United Kingdom, the foundational need is the establishment of routine mental health prevalence data collection and publication within the correctional system, integrating the clinical data that the NHS already holds with the demographic data published by the Ministry of Justice.

## Limitations

Several limitations should be acknowledged. All analyses were conducted on aggregate, population-level data; no individual-level linkage between cognitive test scores, diagnoses, and offence types was possible. The four countries differ in assessment methods (clinician-based versus self-report), target populations (newly admitted versus total population versus sample), time periods, and definitions, making direct prevalence comparisons inappropriate. The CAPAS is a work aptitude assessment, not an intelligence test, and no English-language validity studies exist; WAS scores should be interpreted as descriptive indicators, not clinical measurements. The self-report paradox is consistent with a measurement artefact produced by cognitive impairment, but alternative explanations — including healthy survivor effects, cohort differences in mental health literacy, and differential incarceration patterns — cannot be excluded with aggregate data. Finally, this study examines four high-income countries with relatively well-resourced correctional systems; the findings may not generalise to low- and middle-income settings where data infrastructure is even more limited.

## Conclusion

This study demonstrates that three distinct mechanisms — self-report paradox, classification gaps, and data absence — converge to produce the same outcome across four high-income countries: the statistical invisibility of dementia in correctional populations. These are not failures of clinical knowledge; they are consequences of how data systems were designed. Recognising this structural blindness is the necessary first step toward building correctional health surveillance that includes, rather than excludes, the cognitive conditions that ageing prisoners increasingly carry.

The absence of dementia from correctional statistics is not a gap waiting to be filled — it is a mirror of how correctional systems have understood the people in their care. Designing dementia into the categories is not a technical fix. It is a decision about who counts as ill.

## Data Availability

All data used in this study are publicly available. Japan: e-Stat Correctional Statistics Annual (Ministry of Justice), https://www.e-stat.go.jp/. United States: Bureau of Justice Statistics Survey of Prison Inmates 2016, https://bjs.ojp.gov/. United Kingdom: Ministry of Justice Offender Management Statistics Quarterly, https://www.gov.uk/government/collections/offender-management-statistics-quarterly. Australia: Australian Institute of Health and Welfare National Prisoner Health Data Collection, https://www.aihw.gov.au/; Australian Bureau of Statistics Prisoners in Australia, https://www.abs.gov.au/. No individual-level data were accessed.

https://bjs.ojp.gov/

https://www.aihw.gov.au/

https://www.abs.gov.au/

## Declarations

### Data Availability

All data used in this study are publicly available:

- Japan: e-Stat Correctional Statistics Annual (Ministry of Justice). https://www.e-stat.go.jp/
- United States: Bureau of Justice Statistics Survey of Prison Inmates 2016. https://bjs.ojp.gov/data-collection/survey-prison-inmates-spi
- United Kingdom: Ministry of Justice Offender Management Statistics Quarterly. https://www.gov.uk/government/collections/offender-management-statistics-quarterly
- Australia: Australian Institute of Health and Welfare National Prisoner Health Data Collection. https://www.aihw.gov.au/
- Australian Bureau of Statistics Prisoners in Australia. https://www.abs.gov.au/

No individual-level data were accessed. All analyses used publicly available aggregate statistics. No data sharing agreement is required.

### Ethics Statement

This study involved secondary analysis of de-identified, aggregate-level administrative data published by government agencies. No individual-level data were accessed and no contact with research participants occurred. Institutional ethics review was not required.

### Author Contributions

H.F.: Conceptualization, data curation, formal analysis, visualization, writing (original draft, review and editing).

## Acknowledgements

The author thanks the Ministry of Justice of Japan for maintaining and publicly releasing the Correctional Statistics Annual via e-Stat, and the Bureau of Justice Statistics, AIHW, and UK Ministry of Justice for their publicly available correctional data.

